# Rare Variant Analyses in Ancestrally Diverse Cohorts Reveal Novel ADHD Risk Genes

**DOI:** 10.1101/2025.01.14.25320294

**Authors:** Seulgi Jung, Madison Caballero, Emily Olfson, Jeffrey H. Newcorn, Thomas V. Fernandez, Behrang Mahjani

## Abstract

Attention-deficit/hyperactivity disorder (ADHD) is a highly heritable neurodevelopmental disorder, but its genetic architecture remains incompletely characterized. Rare coding variants, which can profoundly impact gene function, represent an underexplored dimension of ADHD risk. In this study, we analyzed large-scale DNA sequencing datasets from ancestrally diverse cohorts and observed significant enrichment of rare protein-truncating and deleterious missense variants in highly evolutionarily constrained genes. This analysis identified 15 high-confidence ADHD risk genes, including the previously implicated *KDM5B*. Integrating these findings with genome-wide association study (GWAS) data revealed nine enriched pathways, with strong involvement in synapse organization, neuronal development, and chromatin regulation. Protein–protein interaction analyses identified chromatin regulators as central network hubs, and single-cell transcriptomic profiling confirmed their expression in neurons and glial cells, with distinct patterns in oligodendrocyte subtypes. These findings advance our understanding of the genetic architecture of ADHD, uncover core molecular mechanisms, and provide promising directions for future therapeutic development.

## Introduction

Attention-deficit/hyperactivity disorder (ADHD) is a common neurodevelopmental disorder characterized by inattention, hyperactivity, and impulsivity. Its global prevalence ranges from approximately 5.6 to 11.4% in children and adolescents,^1–5^ depending on diagnostic criteria, with at least 2.6% of adults worldwide experiencing persistent symptoms.^6^ While the high prevalence and significant impact of ADHD are well appreciated, its biological underpinnings remain only partially understood. In general, ADHD is widely recognized as arising from a combination of genetic predispositions and environmental influences,^7,8^ with emerging research offering insights into its neurobiological basis.^9^

ADHD exhibits high heritability, with estimates between 70 and 80%, based on findings from family and twin studies.^7^ Genome-wide association studies (GWAS) have identified numerous common genetic markers associated with ADHD, many of which are located within genes influencing neurotransmitter systems crucial for regulating attention and behavioral regulation.^10,11^ These findings emphasize the polygenic nature of the disorder, with multiple genetic variants of small effects collectively contributing to risk. However, the heritability explained by single-nucleotide polymorphisms (SNPs) is only between 14% and 22%, leaving a large proportion unexplained. This discrepancy, often referred to as “missing heritability,“^12^ likely reflects a combination of factors, such as gene-environment interactions, epigenetics, structural variations, and rare genetic variants.

Rare variants, which are not well-captured by GWAS due to their low population frequency, often have larger individual effect sizes and are more likely to disrupt gene function. Whole-exome sequencing (WES) studies have revealed a significant enrichment of rare damaging variants in individuals with ADHD compared to unaffected controls.^13,14^ Notably, lysine demethylase 5B (*KDM5B*) has recently emerged as a high-confidence risk gene, marking an important advance in understanding the genetic basis of ADHD.^14^ Additionally, rare structural alterations in the genome, such as copy number variants (CNVs), have also been implicated as contributors to ADHD risk.^15–18^ Taken together, these findings underscore the pivotal role of rare variants, particularly those that disrupt gene function, in the genetic architecture of ADHD. Unlike common variants, which typically exert small, cumulative effects, rare variants often result in substantial disruptions to protein function, gene regulation, or cellular pathways, providing more direct insights into the biological mechanisms underlying ADHD. Investigating these variants offers an opportunity to uncover key genes and pathways, facilitating the discovery of novel molecular mechanisms and potential therapeutic targets for the disorder.

This study investigates the contribution of rare genetic variants to ADHD risk by analyzing whole-genome sequencing (WGS) and WES data from multiple cohorts with diverse genetic ancestries. By identifying deleterious rare variants, we aim to uncover ADHD risk genes and elucidate the biological pathways enriched in these genes. Additionally, we explore the heterogeneity of ADHD risk genes compared to those implicated in autism spectrum disorder (ASD), offering insights into both shared and distinct genetic architectures underlying these neurodevelopmental disorders.

## Results

We analyzed WGS and WES datasets derived from multiple sources, including the *All of Us* Research Program, the SPARK initiative, and two previously published studies (Olfson et al., 2024; Satterstrom et al., 2019)(Supplementary Table 1).^13,14,19,20^ The datasets included (1) 4,854 ADHD cases (defined using ICD-10 criteria) and 24,108 ancestry- and sex-matched controls from the *All of Us* Research Program. (2) 902 affected trios (self-reported ADHD) and 3,508 unaffected trios from the SPARK initiative. Because SPARK is ascertained for autism spectrum disorder (ASD), individuals with an ASD diagnosis were excluded, and only those with self-reported ADHD were retained as affected trios. Similarly, unaffected trios were defined as individuals without a diagnosis of ASD, ADHD, or any other mental disorders. (3) 3,477 ADHD cases (defined using ICD criteria) and 5,002 controls from Satterstrom et al. (2019). This dataset contains individuals with co-occurring ADHD and ASD as well as those with ADHD alone; we included only individuals with ADHD (defined using ICD criteria) without ASD in our analysis. (4) 147 affected trios (clinically diagnosed ADHD) and 780 unaffected trios from Olfson et al. (2024).

In total, we included 1,049 affected trios, 4,288 unaffected trios, 8,333 cases, and 29,110 controls in our analysis. To focus on rare coding variants, we restricted our analysis to variants located in exons with an allele frequency of ≤0.1% for de novo variants and an allele count ≤ 5 for case-control variants in both our dataset and the non-psychiatric subsets of gnomAD v3.1.^21^

### Burden of rare coding variation

Following the assembly and quality control of the datasets, we compared the burden of rare coding variants between affected and unaffected individuals. Due to their high potential functional impact, these variants may play a critical role in the genetic architecture of ADHD. Specifically, we assessed the impact of rare deleterious coding variants on ADHD risk by analyzing protein-truncating variants (PTVs) and missense variants across all study cohorts.

We categorized genes into deciles based on *loss-of-function observed/expected upper bound fraction* (LOEUF) scores (from 0 to 2), with lower scores reflecting greater selective pressure against PTVs. To ensure sufficient statistical power, we combined deciles 1–3 because deciles 1 and 2 alone contained too few variants for meaningful testing. Excluding the SPARK dataset, we observed significant enrichment of PTVs among ADHD cases in genes within the first-to-third deciles in both trios and case-control cohorts, with this enrichment diminishing as selective constraint decreased (Fig. 1 and Extended Data Fig. 1a-d).

**Fig. 1:**
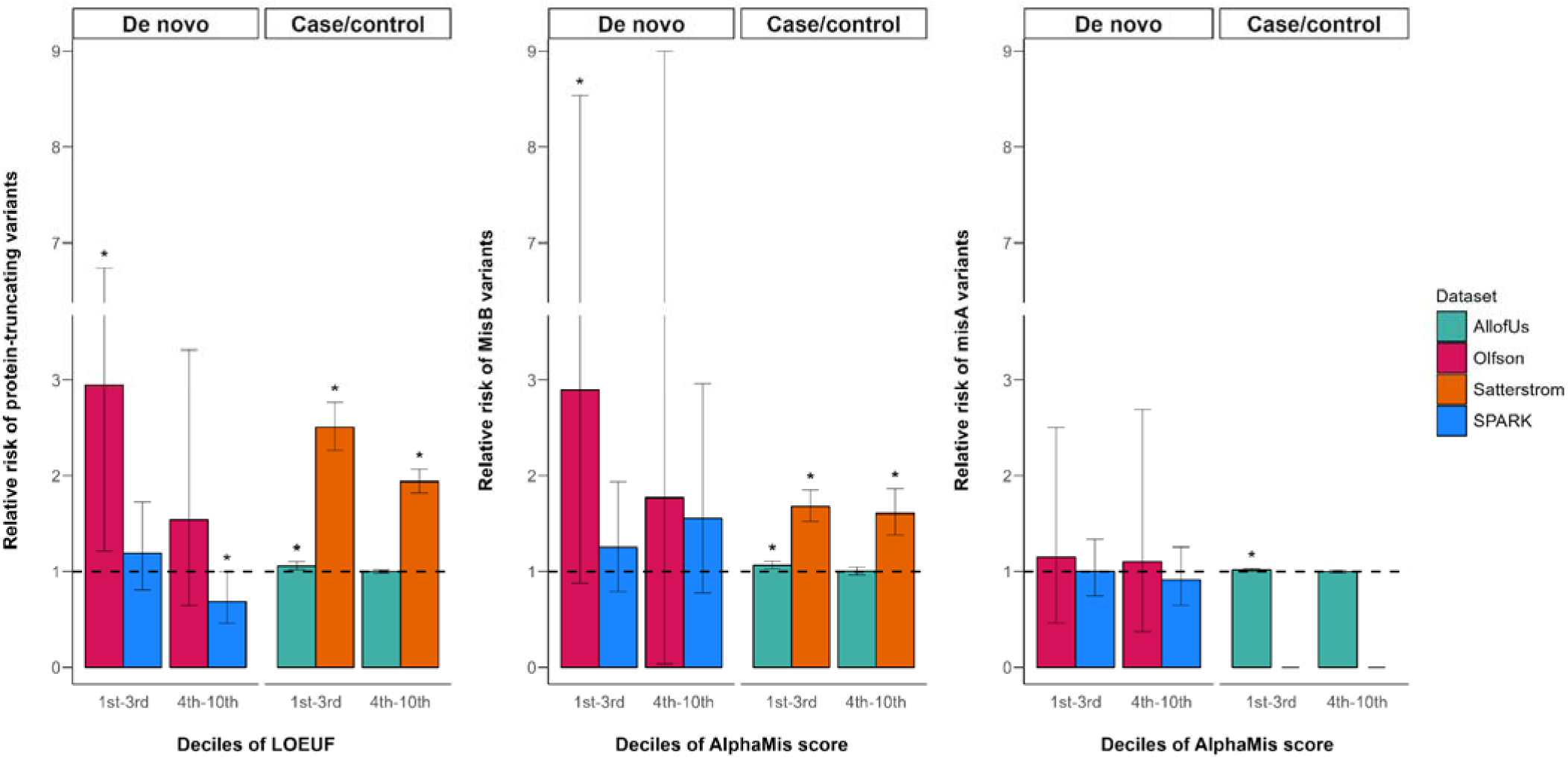
Enrichment of rare autosomal protein-coding variants in ADHD. Frequencies of protein-truncating variants (PTVs) and deleterious missense variants in ADHD case and unaffected control cohorts across LOEUF (Loss-of-function observed/expected upper bound fraction) and AlphaMissense score deciles. Missense variants were classified based on MPC (Missense badness, PolyPhen-2, and Constraint) scores into MisB (MPC ≥ 2) and MisA (2 > MPC ≥ 1) categories. MisA variants were not available in the Satterstrom dataset, so orange bars are omitted from the last graph. \**P*<0.05 using a binomial test between ADHD cases and unaffected controls.

We categorized deleterious missense variants by categorizing them into two subgroups based on their MPC (Missense badness, PolyPhen-2, and Constraint) scores: MisB (MPC ≥ 2) and MisA (2 > MPC ≥ 1).^22^ For MPC, higher scores indicate a greater probability of a variant being pathogenic. Additionally, we stratified genes into deciles according to their AlphaMissense scores, which range from 0 to 1, with higher scores indicating stronger selective pressure against missense variants.^23^ Genes in the top three deciles of AlphaMissense score exhibited significant enrichment of MisB variants in ADHD cases, in both trios and case-control cohorts, excluding the SPARK dataset (Fig 1). In the Olfson and All of Us datasets, this enrichment gradually increased as selective constraints became stronger, however, it was not observed in the SPARK and Satterstrom datasets (Extended Data Fig. 2a-d).

For MisA variants in genes in the top three deciles of AlphaMissense score, we observed significant enrichment in ADHD cases in the All of Us case-control cohort; however, the enrichment pattern, while aligning consistently with the AlphaMissense score, was weaker than that of MisB variants (Fig. 1 and Extended Data Fig. 3a-d). Synonymous variants did not show any significant enrichment in ADHD cases across trios or case-control cohorts.

These findings suggest that rare coding variants, particularly PTVs and deleterious missense variants in conserved genes, represent important genetic risk factors for ADHD.

### Number of risk genes

The observed enrichment of PTV and deleterious missense variants in conserved genes among ADHD cases suggests that these genes are likely to harbor specific genetic risk factors for the disorder. We applied the “unseen species problem” method to estimate the expected number of risk genes that could be identified through rare variant analysis.^24^ This approach, previously used in similar analyses of rare variant contributions to complex traits, infers the expected number of risk-associated genes, including those not yet observed, by analyzing the frequencies of deleterious *de novo* variants and estimating the likelihood that these variants represent a subset of the complete set of risk genes.

Given the SPARK dataset is ascertained for ASD, and parental data are required to distinguish inherited variants from de novo variants, we restricted our analysis to the trios from the Olfson et al.(2024) dataset to minimize potential bias arising from ASD-specific ascertainment. Using this cohort, we examined the number of deleterious de novo PTVs and missense variants predicted to be damaging (MisB) (Methods).

Our analysis estimated approximately 173 autosomal coding genes, representing 0.95% of the 18,128 protein-coding genes analyzed, as potentially implicated in ADHD through rare deleterious variation. However, these estimates are provisional due to the limited sample size and should be refined as additional data becomes available.

### Identification of risk genes

We next sought to identify specific risk genes for ADHD. The relative risk associated with ADHD-related variants can vary based on factors such as variant type (de novo and case/control), variant classification (PTV, MisB, and MisA), and evolutionary constraint (LOEUF and AlphaMis scores). Integrating these factors in association testing can improve gene prioritization and enhance power to detect true associations.

To leverage this variation and identify ADHD risk genes, we applied the TADA^25^, an approach that integrates rare deleterious variants across various classes while accounting for evolutionary constraints, mutation rates, and prior probabilities of relative risk, as well as variant types and inheritance patterns. For each autosomal protein-coding gene, we calculated a Bayes factor (BF) and false discovery rate (FDR) to quantify the strength of evidence for an association (Supplementary Table 2). In accordance with the All of Us Data and Statistics Dissemination Policy, we are unable to display the ultra-rare variant counts per gene.

Because the SPARK dataset is ascertained for ASD, we used the Olfson et al.(2024) dataset for de novo variants and the All of Us dataset for case-control variants to estimate TADA parameters and prior probabilities of relative risk, thus minimizing ASD-specific ascertainment bias.

We assessed potential inflation in TADA results by calculating the genomic inflation factor (λ_GC_) and examining a quantile-quantile (Q-Q) plot. The genomic inflation factor was 1.017, indicating minimal inflation and suggesting that observed p-values were not significantly confounded by population stratification or other biases. The Q-Q plot corroborated this finding, with observed p-values closely aligning with the expected diagonal, apart from upward deviations for some genes, indicative of true associations (Extended Data Fig. 4).

Our analysis identified 15 genes with a q value (FDR) less than 0.1, a threshold commonly used in rare variant studies to identify risk genes and minimize false discoveries.^14,25–28^ Among these were *KDM5B*, a histone demethylase enzyme involved in regulating gene transcription, cell differentiation, and proliferation,^29^ previously implicated as an ADHD risk gene by Olfson et al. (2024), and 14 novel risk genes (Table 1 and Fig. 2).

**Fig. 2:**
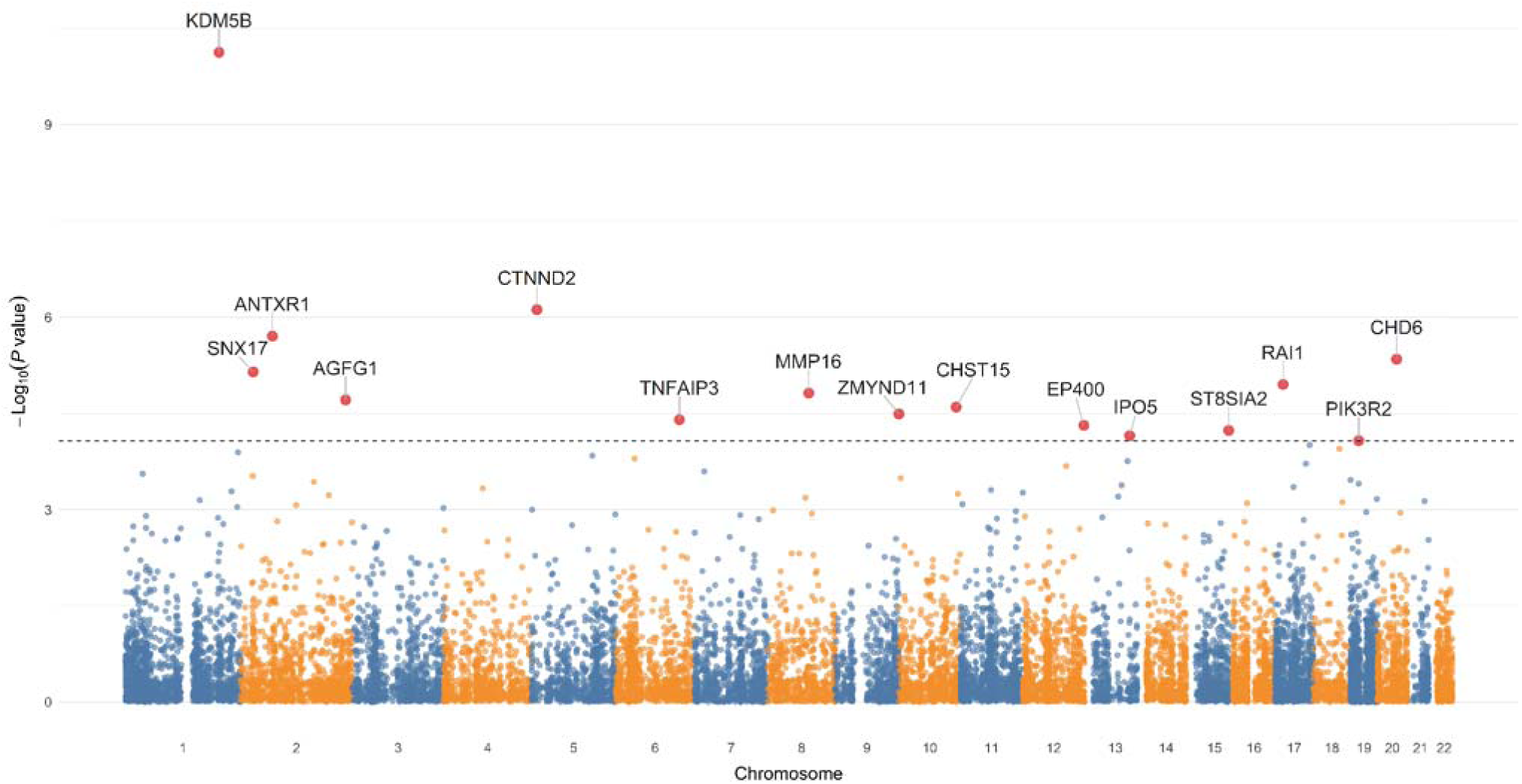
Association of protein-coding genes with ADHD risk. Each point represents the - log_10_(*P* value) for the association of individual protein-coding genes in the ADHD TADA result. Genes with a q value < 0.1 (red points and dotted line) are labeled.

**Table 1.** Genes with q value <0.1 in TADA result.

| Gene | LOEUF | pLI | q value<sup>*</sup> | P value<sup>#</sup> |
| --- | --- | --- | --- | --- |
| *KDM5B* | 0.572 | 0.000 | 0.000 | $7.47 \times 10^{-11}$ |
| *CTNND2* | 0.098 | 1.000 | 0.007 | $7.64 \times 10^{-7}$ |
| *ANTXR1* | 0.3 | 0.986 | 0.012 | $1.96 \times 10^{-6}$ |
| *CHD6* | 0.073 | 1.000 | 0.020 | $4.50 \times 10^{-6}$ |
| *SNX17* | 0.579 | 0.002 | 0.025 | $7.08 \times 10^{-6}$ |
| *RAI1* | 0.116 | 1.000 | 0.033 | $1.12 \times 10^{-5}$ |
| *MMP16* | 0.35 | 0.918 | 0.039 | $1.53 \times 10^{-5}$ |
| *AGFG1* | 0.278 | 0.993 | 0.043 | $1.94 \times 10^{-5}$ |
| *CHST15* | 0.803 | 0.000 | 0.050 | $2.52 \times 10^{-5}$ |
| *ZMYND11* | 0.12 | 1.000 | 0.057 | $3.22 \times 10^{-5}$ |
| *TNFAIP3* | 0.196 | 1.000 | 0.064 | $3.96 \times 10^{-5}$ |
| *EP400* | 0.208 | 1.000 | 0.072 | $4.83 \times 10^{-5}$ |
| *ST8SIA2* | 0.651 | 0.058 | 0.079 | $5.79 \times 10^{-5}$ |
| *IPO5* | 0.122 | 1.000 | 0.089 | $7.00 \times 10^{-5}$ |
| *PIK3R2* | 0.489 | 0.016 | 0.099 | $8.37 \times 10^{-5}$ |

LOEUF, loss-of-function observed/expected upper bound fraction score; pLI, the probability of being loss-of-function intolerant score; TADA, transmission and de novo association.
<sup>\*</sup>q value: false discovery rate, calculated in transmission and de novo association test.
<sup>#</sup>P value: P value of transmission and de novo association test.

As sensitivity analyses, we re-applied TADA after excluding the SPARK cohort to minimize the influence of ASD-specific ascertainment. In this analysis, six genes (*KDM5B*, *CTNND2*, *MMP16*, *RAI1*, *ZMYND11*, and *ST8SIA2*) of the originally identified 15 genes remained significant at q value < 0.1 (Supplementary Table 3). Furthermore, when TADA was restricted solely to the two case-control datasets (All of Us and Satterstrom), two genes (RAI1 and ZMYND11) continued to meet q value < 0.1 (Supplementary Table 4). These findings suggest that the associations for at least a subset of the identified genes are not wholly dependent on the inclusion of the SPARK cohort or signals from the de novo variants.

#### Expression pattern of ADHD risk genes

We next examined the expression profiles of the 15 identified ADHD risk genes using publicly available RNA sequencing datasets from the Gene Expression Omnibus (GEO; http://www.ncbi.nlm.nih.gov/geo/).^30^ Among the available data, we identified a whole-blood RNA-seq study comprising both monozygotic twin pairs and case-control samples with and without ADHD.^31^ Differential expression analyses were performed on 16 probands and their unaffected siblings, as well as on 23 cases and 21 controls.

In the twin dataset, *CTNND2* (catenin delta 2) expression was significantly reduced in ADHD probands compared to their siblings. In the case-control dataset, *ST8SIA2* (ST8 alpha-N-acetyl-neuraminide alpha-2,8-sialyltransferase 2) and *ANTXR1* (ANTXR cell adhesion molecule 1) were significantly downregulated in ADHD cases compared to controls (log fold change > |1|, *P* < 0.05) (Supplementary Table 5). Although these findings did not remain significant after correction for multiple testing (FDR > 0.1), The observed differentially reduced expression in unrelated ADHD probands may still indicate that certain rare deleterious variants disrupt protein production, potentially affecting critical cellular processes and contributing to ADHD pathophysiology.

To further investigate the cell type-specific expression patterns of the 15 ADHD risk genes, we leveraged the cellxgene platform (https://cellxgene.cziscience.com/gene-expression) to analyze single-cell RNA sequencing data.^32^ All 15 ADHD risk genes were expressed across various neural cell types, including both neurons and glial cells (Extended Data Fig. 5).

#### Pathway Enrichment Analysis

To determine whether ADHD risk genes are overrepresented in specific biological pathways, we performed pathway enrichment analysis using the GENE2FUNC module from the Functional Mapping and Annotation (FUMA) platform.^33^ The analysis included BioCarta, KEGG, Reactome pathways, and Gene Ontology (GO) terms. Multiple testing correction was applied using the Benjamini–Hochberg method (adjusted *P* < 0.05). Initial analysis of the 15 risk genes identified in our rare variant study did not reveal significant enrichment in any pathways.

To expand the list of ADHD risk genes, we performed gene-based annotation using MAGMA (Multi-marker Analysis of Genomic Annotation) with the summary statistics from the current ADHD GWAS.^11,34^ MAGMA annotated a total of 42 genes with significant *P* values after Bonferroni correction (Supplementary Table 6). Using a combined set of 57 genes (15 from the rare variant study and 42 from GWAS), we identified 9 Gene Ontology biological pathways with FDR<0.05, including ‘synapse organization’ and ‘generation of neurons’ (Supplementary Table 7). All nine pathways included at least one gene from our rare variant study. Notably, the ‘developmental growth’ and ‘cell morphogenesis’ pathways emerged only after incorporating the 15 genes from the rare variant study.

#### Protein-protein Interaction Analysis

We next used the Search Tool for the Retrieval of Interacting Genes/Proteins (STRING) to examine protein-protein interactions (PPI) among the 15 ADHD risk genes identified from our rare variant analysis.^35^ Unlike pathway enrichment analysis, which focuses on the functional annotation of genes in broader biological contexts, STRING highlights PPI and the functional connectivity between encoded proteins. Among the 15 ADHD risk genes, *KDM5B, CHD6, ZMYND11, and EP400* emerged as central hubs within the resulting PPI network. These genes were connected and associated with the ‘chromatin regulator’ biological process (FDR<0.05) (Extended Data Fig. 6a and Supplementary Table 8).

We then performed a PPI analysis on the combined set of 57 ADHD-associated genes (15 from the rare variant study and 42 from GWAS). This resulted in a PPI network comprising 160 edges, significantly more than the 79 edges expected by chance (PPI enrichment *P* value = 1.1 × 10^−15^). These findings indicate functionally interconnected among the input genes (Extended Data Fig. 6b). Functional enrichment analysis within this expanded network highlighted significant involvement in pathways such as ‘regulation of synapse organization’ (FDR<0.05) (Extended Data Fig. 6b and Supplementary Table 8). These results complement the pathway enrichment analysis using the GENE2FUNC module of FUMA above by identifying proteins that directly interact within the highlighted pathways, underscoring the functional roles of genes like *ST8SIA2* as central connectors in synaptic processes.

#### Ancestry Analysis

We identified 15 genes associated with ADHD risk; however, it remains unclear whether the genetic architecture of ADHD, particularly involving rare damaging variants, varies across ancestries. While the fundamental biological constraints on damaging variants are unlikely to differ by ancestry, the occurrence and frequency of such variants may be shaped by historical demographic events, including bottlenecks, founder effects, and genetic drift. To investigate this, we first examined the population frequency of rare synonymous and potentially damaging variants (PTVs, MisB, and MisA) in these genes using data from gnomAD and the *All of Us* Research Program across ancestries [non-Finnish European (NFE), African (AFR), and admixed-American (AMR) populations]. This initial analysis enabled us to distinguish ordinary ancestral variation from potential ADHD-specific genetic signatures. We found that the genetic architecture of these 15 genes, including both potentially neutral (synonymous) and deleterious (PTV, MisB, and MisA) variants, differed markedly between ancestries (Supplementary Table 9). While these findings do not preclude the presence of ADHD-associated rare variants, they underscore the importance of interpreting any observed enrichment of damaging variants within the context of established population-level baselines. Failure to account for such ancestral differences could lead to misattribution of ordinary population variation to disease risk.

We compared the burden of rare damaging variants in the 15 ADHD risk genes among ADHD cases in the All of Us cohort, stratified by ancestry. The test identified *CTNND2* as significantly different (*P* < 0.001) between NFE and AMR populations, and *EP400* as significantly different (*P* < 0.001) between NFE and AFR populations (Supplementary Table 10). Then, we performed the Breslow–Day test to assess whether these differences remained significant after accounting for ancestry-specific baseline frequencies. For *CTNND2*, the Breslow–Day test indicated that baseline differences between ancestries explained the observed signal, as its significance disappeared. For *EP400*, the *P*-value increased to 3.19E-04, suggesting that part of the observed association was due to population-level variation (Supplementary Table 10). However, the association remained statistically significant, indicating that differences in variant frequencies for *EP400* persist even after adjustment.

In AFR, the burden of ultra-rare variants in *EP400* was higher (0.02155), and the associated OR > 1 (1.627) suggests a notable contribution of this gene to ADHD risk in this population. In contrast, in NFE, the burden of ultra-rare variants was much lower (0.00319), and the OR < 1 (0.575) likely reflects the reduced contribution of *EP400* to ADHD in this population. This finding does not imply that *EP400* is biologically protective in NFE but rather that deleterious variants in this gene are less frequently observed and may not be a major contributor to ADHD risk in European-ancestry populations. However, these findings should be approached with caution due to the limited sample size for rare variant analyses.

#### Relationship between ADHD and ASD

Of the 15 ADHD risk genes identified through our rare variant analysis, we found that three genes have also been implicated as risk genes for ASD: *KDM5B*, *RAI1* (retinoic acid induced 1), and *ZMYND11* (zinc finger MYND domain-containing protein 11).^25^ To investigate whether the burden of deleterious rare coding variants differs between ADHD and ASD, we performed heterogeneity analyses using de novo and case-control variant data from 15,036 ASD probands and 5,591 ASD cases.^25^

Finally, we counted PTVs, MisB, and MisA variants within the 15 ADHD risk genes and the remaining genes (18,128 genes excluding the 15). We performed heterogeneity analyses using Fisher’s exact test between ADHD and ASD (Supplementary Table 11). Deleterious de novo variants were significantly more enriched in ADHD probands compared to ASD probands in four genes: *CHST15* (carbohydrate sulfotransferase 15), *CHD6* (chromodomain helicase DNA binding protein 6), *CTNND2*, and *TNFAIP3* (tumor necrosis factor alpha-induced protein 3) (FDR<0.05). Five additional genes (*EP400*, *AGFG1*, *ANTXR1*, *MMP16*, and *SNX17*) showed nominally significant heterogeneity in variant enrichment.

## Discussion

Our study aimed to elucidate the genetic risk architecture of ADHD through an in-depth analysis of rare coding variants. We observed significant enrichment of PTVs and deleterious missense variants in genes under strong evolutionary constraint, as reflected by top decile rankings in LOEUF and AlphaMissense scores among individuals with ADHD. Building on these findings, we identified 14 novel risk genes for ADHD and replicated previous findings associated with *KDM5B*. We identified additional de novo PTV in the SPARK dataset and significant enrichment of PTVs and deleterious missense variants in case-control analyses in *KDM5B*. *KDM5B* plays a vital role in regulating gene expression and synaptic plasticity in the adult hippocampus.^36^

The second significant gene, *CTNND2*, encodes delta-catenin, which is expressed in proliferating neuronal progenitor cells of the neuroepithelium and the dendritic compartments of postmitotic neurons.^37^ *CTNND2* plays a critical role in regulating synaptic maturation, neural excitability, and the accumulation of SYNGAP1, with SRGAP2 proteins modulating its synaptic function and contributing to human synaptic neoteny.^38^ A study has implicated in *CTNND2* in ADHD pathophysiology through chromosomal translocation and disruption, linking it to severe ADHD phenotypes and potential genetic mechanisms.^39^

Haploinsufficiency of *RAI1*, which encodes a nuclear protein, leads to Smith-Magenis syndrome, a condition presenting a range of neurodevelopmental and behavioral characteristics.^40,41^ *RAI1* predominantly associates with DNA regions proximal to active promoters, enhancing the transcription of genes involved in neural circuit formation and neuronal communication.^42^

*MMP16*, a member of the matrix metalloproteinase which is involved in various physiological and pathological processes, including morphogenesis, wound healing, tissue repair, and tissue remodeling, was identified as a genetic locus associated with schizophrenia.^43^ Furthermore, rare copy-number variation in *MMP16* was observed in individuals with ADHD.^44^

The role of *EP400*, a critical chromatin remodeling factor, in regulating gene expression relevant to neuronal development and synaptic function has been identified as a causative factor for epilepsy and neurodevelopmental disorders.^45^

Overall, these results suggest that genes under evolutionary constraint are more likely to harbor rare variants that impact ADHD risk, emphasizing their critical importance as targets for genetic and functional research. This observation aligns with prior research implicating constrained genes in the pathogenesis of various psychiatric disorders.^25–28,46–48^ Interestingly, a few risk genes like *KDM5B*, *SNX17*, *CHST15*, *ST8SIA2* and *PIK3R2* exhibited less constrained LOEUF scores compared to other genes, highlighting the need to investigate how these types of genes contribute to ADHD pathophysiology.

Gene expression analyses in unrelated ADHD probands further revealed reduced expression of several risk genes identified by rare variants, including *CTNND2*, *ST8SIA2*, and *ANTXR1*, pointing to potential disruptions in protein production and cellular function. To gain deeper insights into how these genes function within specific cell types, we analyzed single-cell RNA sequencing data using the CellxGene platform. We found that all 15 ADHD risk genes that we identified were expressed in both neurons and glial cells, indicating their broad involvement in brain function. Additionally, transcriptomic profiles from adult human brain tissue revealed that five ADHD risk genes (*CTNND2, MMP16, AGFG1, RAI1*, and *EP400*) display distinct expression patterns across two oligodendrocyte subtypes—OPALIN+ and RBFOX1+ cells—that differ in maturation state.^49^ Notably, *CTNND2*, *MMP16, RAI1*, and *EP400* were enriched in OPALIN+ oligodendrocytes associated with active myelination, while *AGFG1* was more highly expressed in RBFOX1+ oligodendrocytes indicative of a more mature lineage. Oligodendrocytes play a pivotal role in cognitive function.^50,51^ The evolutionary changes in oligodendrocyte gene expression may influence cognitive disorders, including ADHD.^52^ These findings suggest that these genes may influence oligodendrocyte differentiation and function, potentially shaping the development and maintenance of neural circuits implicated in ADHD.

To further explore the biological implications of these findings, we conducted pathway enrichment and PPI analyses. These complementary methods provided a more comprehensive view of the underlying biological mechanisms. Pathway enrichment analysis primarily highlights functionally enriched sets of genes within known biological pathways, while PPI analysis uncovers the physical and functional interactions among proteins encoded by these genes, revealing how they coalesce into interconnected networks. By integrating these approaches, our study identified core developmental processes, such as neurogenesis, neuronal differentiation, synapse organization, regulation of synapse activity, and chromatin regulation that are closely linked to the ADHD risk genes that we identified.

These core processes encompass essential aspects of brain organization and function, offering insights into how disruptions in these pathways may contribute to ADHD. Developmental and neuron-related pathways form the foundation for brain organization, encompassing neurogenesis, neuronal specialization, and the formation of functional networks critical for cognitive and behavioral regulation. Disruptions in these processes are often implicated in ADHD.^53–56^ Impairments in synaptic organization and adaptability may disrupt neurotransmitter signaling, limiting the brain’s capacity for flexible responses in individuals with ADHD.^57–59^ Furthermore, chromatin regulation integrates epigenetic processes that modulate gene expression during brain development. Dysregulation in these pathways may result in improper activation or silencing of genes essential for neural function, thereby contributing to ADHD pathophysiology.^60–62^ Through the integration of pathway enrichment and PPI analyses, we identified enriched biological processes while uncovering the molecular architecture and interconnectedness of gene networks that underpin the pathophysiology of ADHD.

The substantial involvement of chromatin regulation and developmental pathways in ADHD raises intriguing questions about its genetic and neurobiological overlap with ASD. Studies have shown that 15% to 25% of children with ADHD exhibit traits and symptoms of ASD, with 12.4% meeting the diagnostic criteria for ASD.^63–65^ Conversely, ADHD is one of the most frequently observed comorbidities in children with ASD, with reported rates ranging from 40% to 70%.^66–69^ This significant clinical overlap highlights the potential for shared genetic mechanisms between these conditions. Supporting this hypothesis, a previous study has demonstrated a similar burden of rare PTVs in evolutionarily constrained genes among individuals with ADHD and ASD.^13^ Among 15 ADHD risk genes we identified, *KDM5B*, *RAI1*, and *ZMYND11* have also been reported as ASD risk genes.^25^ Notably, these genes are involved in transcriptional regulation via chromatin modification,^70–74^ suggesting that disruptions in these processes may contribute to the co-occurrence of ASD and ADHD.

At the same time, four genes (*CHST15*, *CHD6*, *CTNND2*, and *TNFAIP3*) exhibited significant enrichment for deleterious rare de novo variants in ADHD compared to ASD, suggesting disorder-specific genetic contributions. Of particular interest, *TNFAIP3*, an immune-related gene, serves as a negative regulator of TLR immune response pathways and has been associated with major depressive disorder.^75–77^ Loss of one copy of *TNFAIP3* (A20) can lead to cognitive symptoms and neuroinflammation,^78^ suggesting potential immune system involvement in ADHD pathophysiology.^79^ These findings indicate that while ADHD and ASD share genetic risk factors, they also exhibit distinct genetic profiles.

Our analysis of ultra-rare damaging variants in 15 ADHD-associated genes revealed ancestry-specific differences in *EP400*. In AFR populations, *EP400* showed a higher burden of ultra-rare variants (0.02155) and an OR > 1 (1.627), indicating a significant contribution to ADHD risk. In contrast, the much lower burden in NFE (0.00319) and OR < 1 (0.575) likely reflects a reduced role of this gene in ADHD risk rather than true biological protection. This finding should be interpreted cautiously, as low variant counts and limited sample sizes may produce statistical artifacts. These results highlight the importance of ancestry-specific analyses in understanding rare variants’ contributions to ADHD risk.

This study has several notable strengths. One of its key strengths is the integration of rare variant analyses with functional data, including gene expression and pathway-level insights. This approach enabled the identification of both shared and distinct genetic mechanisms underlying ADHD and ASD. Additionally, the use of complementary methods, such as single-cell RNA sequencing and transcriptomic profiling, allowed for a detailed exploration of the cellular contexts in which these risk genes are expressed, shedding light on their potential roles in neurodevelopment. Together, these strategies provided a comprehensive framework for understanding the genetic and molecular basis of ADHD.

However, this study also has limitations. One major limitation is the lack of experimental validation for the identified risk genes, which leaves uncertainty about their precise functional roles. Future research should prioritize functional assays, such as CRISPR-based models or other in vitro and in vivo approaches, to clarify the biological effects of these variants. Additionally, while the study identified key pathways involved in ADHD, the broader interactions between genetic, epigenetic, and environmental factors remain unexplored. Another notable limitation is the heterogeneity of ADHD itself. By categorizing all participants under a single ADHD diagnosis, this study may have overlooked genetic contributions specific to subtypes or symptom dimensions, such as inattentiveness, hyperactivity, or impulsivity. Future research should aim to integrate genetic insights with detailed clinical phenotyping to address this heterogeneity, enabling a more nuanced understanding of how genetic factors contribute to distinct presentations of ADHD. Additionally, a notable limitation of our study is the use of unaffected siblings from ASD trios in the SSC and SPARK cohorts as controls. While these individuals are considered unaffected by ASD, it is important to acknowledge that both cohorts are derived from ASD-enriched populations, which may introduce potential biases. Moreover, there are differences in the depth of phenotypic assessment for psychiatric disorders between these cohorts. While diagnoses in the SPARK cohort were based on self-reports, the SSC cohort, as used in the Olfson cohort, underwent more thorough evaluations for psychiatric disorders. Consequently, we utilized the Olfson cohort to calculate the number of risk genes and parameters for TADA. By using these controls in an ADHD-focused study, we acknowledge that this limitation could bias the results toward the null hypothesis, potentially reducing the ability to detect significant associations.

In summary, this study provides critical insights into the genetic architecture of ADHD, identifying 14 novel risk genes and uncovering key biological pathways, including chromatin regulation, neurogenesis, and synapse organization, that contribute to its pathophysiology. Our findings demonstrate both shared and distinct genetic contributions with ASD, highlighting the overlapping yet divergent molecular mechanisms underlying these neurodevelopmental disorders. Importantly, the identification of chromatin-regulating genes and immune-related factors, such as *TNFAIP3*, points to novel avenues for therapeutic exploration. Understanding how these pathways interact may enable precision medicine approaches, facilitating targeted interventions for individuals with ADHD, particularly those with overlapping features of ASD or immune dysregulation. Future research should integrate genetic insights with detailed clinical phenotyping to better predict outcomes and tailor treatment strategies, ultimately improving care for individuals affected by ADHD and related neurodevelopmental conditions.

## Methods

### Study population: All of Us

The All of Us Research Program, led by the National Institutes of Health (NIH), is a landmark precision medicine initiative aimed at accelerating health research and medical breakthroughs by gathering a diverse set of data from one million or more participants across the United States.^20^ In the current All of Us Research Program (controlled tier, version 7) workspace, we identified 7,198 samples diagnosed with attention-deficit/hyperactivity disorder (ADHD). Of these, 5,063 samples had available whole-genome sequencing (WGS) data. For our case-control matching analysis, we selected these 5,063 ADHD samples and 153,678 samples without mental disorders (F01-F99 defined using ICD-10 criteria) from the WGS dataset. Using the R package optmatch, we performed 1:5 case-control matching based on sex and principal components (PCs) 1–5, which were calculated from the WGS data and provided by the All of Us Research Program. We confirmed that the resulting 5,063 cases and 25,315 controls were well matched, as shown by scatter plots (Extended Data Fig. 7).

### Study population: SPARK

The Simons Foundation Powering Autism Research for Knowledge (SPARK) project is a large-scale research initiative designed to enhance our understanding of autism spectrum disorder (ASD) by building a comprehensive database containing genetic, behavioral, and medical information from individuals with ASD and their families.^19^ The third data release (August 2024) from SPARK, which integrated whole exome sequencing (WES), included 142,357 samples, comprising 40,193 complete trios—28,159 with ASD and 12,034 unaffected trios. Among the unaffected trios, 1,241 included a child diagnosed with ADHD. We utilized the remaining 10,793 unaffected trios without a child diagnosed with ADHD as controls. The diagnoses in the SPARK cohort were based on self-reports.

#### Study population: Summary statistics from other studies

We incorporated 150 and 662 rare de novo variants from 147 ADHD trios and 780 unaffected trios, respectively, as reported in Olfson et al., 2024 (Supplementary Table 12).^14^ We used the liftOver tool to convert the genomic positions of the de novo variants to the GRCh38 assembly.^80^ We also used the gene-level rare variant count data from 3,477 cases and 5,002 controls, as reported in Satterstrom et al., 2019.^13^

### Identification of rare coding variants - All of Us

We analyzed short-read whole-genome sequencing (WGS) data from 5,063 cases and 25,315 controls without mental disorders (F01-F99 defined using ICD-10 criteria) obtained from the All of Us Research Program. Following the All of Us guidelines (All of Us Genomic Quality Report [ARCHIVED C2022Q4R9 CDR v7], n.d.), we excluded 43 individuals who had fewer than 2.4 million or more than 5.0 million total variants, over 100,000 variants not present in gnomAD v3.1,^21^ or a heterozygous-to-homozygous variant ratio greater than 3.3.

We removed 1,043,732 low-quality variants defined by genotype quality (GQ) ≤ 20, read depth (DP) ≤ 10, allele balance (AB) ≤ 0.2 for heterozygotes, ExcessHet < 54.69, and quality scores (QUAL) < 60 for single nucleotide variants (SNVs) and < 69 for short insertions and deletions (InDels), as well as variants located in low-complexity regions. Our quality control process further excluded 235 samples with call rates more than three standard deviations below the mean, 426 individuals with close genetic relatedness (kinship coefficient > 0.1) as determined by Kinship-based INference for Genome-wide association studies (KING),^81^ and 710 individuals with sex discrepancies.

After filtering, we retained 4,856 cases and 24,108 controls. We further excluded homozygous reference calls with less than 90% read depth supporting the reference allele or GQ < 25; heterozygous calls with an allele balance < 30%, GQ < 25, or a probability greater than 1×10⁻⁹ for the observed allele balance based on a binomial distribution centered at 0.5; and homozygous alternate calls with less than 90% read depth supporting the alternate allele or GQ < 25. Variants with a call rate ≤ 90% and Hardy-Weinberg equilibrium *P* < 1×10_¹² were also removed. After all filtering steps, 32,687,442 variants were retained.

We utilized the Variant Effect Predictor (VEP) to predict the potential functional effects of the variants.^82^ Variants were classified into protein-truncating variants (PTVs)—including frameshift, stop-gained, splice donor, and splice acceptor variants—missense variants, and synonymous variants. For PTVs, we applied the Loss-Of-Function Transcript Effect Estimator (LOFTEE), requiring a “high-confidence” (HC) designation and excluding LOFTEE flags except “SINGLE_EXON.” Missense variants were categorized based on MPC scores into MisB (MPC ≥ 2), MisA (1 ≤ MPC < 2), and Mis0 (0 ≤ MPC < 1).^22^ To filter rare variants, we included those with an allele count of ≤ 5 in both our case-control cohort and the non-psychiatric subset of the gnomAD database v3.1.^21^

### Identification of rare de novo coding variants - SPARK

The most current WES data (iWES v3) was released in August 2024, consisting of 1-9 batches.^19^ Comprehensive quality control (QC) of batch 5-9 was conducted for both genotypes and samples. For genotype QC, we applied several filters to remove certain genotype calls. We excluded genotype calls with a depth below 10 or above 1,000 and removed genotype calls from the Y chromosome in female samples. Homozygous reference calls were excluded if less than 90% of the read depth supported the reference allele or if the genotype quality (GQ) was less than 25.

Heterozygous calls were removed under several conditions: if the Phred-scaled likelihood of being homozygous reference (PL[HomRef]) < 25; if the call rate (the read depth supporting either the reference or alternate allele) was less than 90%; if the allele balance (the number of mapped reads supporting the alternate allele divided by the read depth) was less than 25%; if the probability of the allele balance, based on a binomial distribution centered on 0.5, was less than 1×10^−9^; or if they were located on the X or Y chromosome (excluding pseudoautosomal regions) of male samples. Homozygous alternate calls were excluded if less than 90% of the read depth supported the alternate allele or if PL[HomRef] < 25.

We also removed samples based on specific criteria. Samples with a high proportion of contaminated reads > 5%. Additionally, we excluded samples with a call rate more than three standard deviations below the mean of each cohort, and duplicated samples. We verified reported pedigrees of family data using the genetic relatedness matrix calculated by KING (kinship coefficient: 0.177-0.354 for 1st-degree relationship).^81^ We removed all samples from incomplete trios. For the final variant QC, we removed variants with a call rate < 10%, or with a Hardy-Weinberg equilibrium (HWE) *P* value < 10^−12^.

The de_novo() function in Hail v0.2.60 (https://hail.is) was used to identify de novo variants from the QC-ed joint-genotyped data (removing variants with a GQ < 25). The population allele frequency of the non-neuro subset from the gnomAD database was used as the input priors of the de_novo() function.^21^ We also used “max_parent_ab = 0.03”, “min_child_ab = 0.3”, and “min_dp_ratio = 0.3” as parameters of the de_novo() function to exclude de novo candidates if the allele balance in a parent is above 3%, if the allele balance of proband is lower than 30%, or if the ratio of read depth between the proband and parents is lower than 30%. This process yielded 214,942 putative de novo variants from a total of 12,887 probands.

Quality Control on De Novo Variants: From the initial 214,942 variants, we kept 56,881 variants with “HIGH” or “MEDIUM” confidence indicated by the calling algorithm (the “MEDIUM” confidence calls are limited to a singleton). We further removed 4,892 and 1,101 variants with allele frequency > 0.1% in the QC-ed joint-genotyped data and non-neuro subset from the gnomAD database, respectively. Then, 4,148 variants were excluded as they appeared more than twice, and an additional 19,567 variants were removed to retain one variant with the most severe effect in the same gene for each sample. Finally, 5,047 variants identified from 49 children with >7 de novo variants were removed. After QC, a total of 22,127 high confidence de novo variants of 12,838 trios remained. After adding the SPARK batch 1–4 data from the existing paper to our dataset,^25^ we retained a total of 9,592 sibling trios without ASD. This combined dataset included 983 unaffected siblings diagnosed with ADHD and 6,331 unaffected siblings without any mental disorders (self-reports from the basic medical screening file). We then excluded all unaffected siblings younger than six years of age, removing 76 siblings with ADHD and 2,812 siblings without any mental disorders. In addition, we removed 16 unaffected siblings who carried more than seven de novo variants (5 with ADHD and 11 without mental disorders). After these steps, the final sample consisted of 902 unaffected siblings with ADHD and 3,508 unaffected siblings without any mental disorders (Supplementary Table 13). High-quality de novo variants from the final sample were listed in Supplementary Table 12.

### Calculating the number of risk genes

We examined the number of deleterious *de novo* variants (PTVs and MisB variants) among 147 ADHD and 780 unaffected trios from the Olfson dataset.^14^ We proceeded under the assumption that the number of deleterious variants in cases minus those in controls (d) contributes to the risk of ADHD, and genes exhibiting recurrent variants (x) were regarded as indicative of risk-associated events. We applied the formula to calculate the number of risk genes in each permutation (C): C = c/u + g^2 * d * (1-u)/u, in which: c as the total number of distinct genes with a *de novo* (d - x), c1 as the count of singleton genes (d - 2x), g as the coefficient of variation in the fractions of genes of each type, and u as 1 – c1/d.^24,83^ For our calculations, we assume g to be 1 due to the limited number of observations.

### Identification of risk genes

We employed the Transmission and De Novo Association Test (TADA) to pinpoint genes associated with ADHD.^84^ We categorized deleterious missense variants by categorizing them into two subgroups based on their MPC (Missense badness, PolyPhen-2, and Constraint) scores: MisB (MPC ≥ 2) and MisA (2 > MPC ≥ 1).^22^ Additionally, we added both *loss-of-function observed/expected upper bound fraction* (LOEUF) and AlphaMissense scores to each gene. Then, we incorporated rare coding variants (PTVs, MisB, MisA, and synonymous variants) to each gene (Supplementary Table 2). Due to the All of Us Data and Statistics Dissemination Policy, we were unable to display the individual ultra-rare variant counts per gene.

The extended version of TADA calculated a Bayes Factor (BF) to measure the statistical strength of the gene-level association.^25,26^ This calculation draws on several inputs, including the number of variant events, sample sizes, the proportion of risk genes, and a prior parameter (gamma) reflecting the variant risk within each gene. For each gene, the gamma values for PTVs, MisB variants, and MisA variants were derived by dividing the variant’s relative risk by the estimated proportion of genes that significantly contribute to ADHD, then smoothing these values over the LOEUF score for PTVs and AlphaMissense score for missense variants. We organized the data into LOEUF- or AlphaMissense-ranked bins, fitted a logistic curve, and calculated a rolling average gamma for PTV, MisB, and MisA variants to represent their relative enrichment in cases versus controls (Supplementary Table 2).

To obtain the total gene-level BF, we multiplied the BFs from the three variant classes. We then imposed a lower bound of 1 for each variant class’s BF to prevent one class from weakening the association evidence provided by another. These BFs were transformed into posterior probabilities, which served as the basis for calculating *P* values and q values for multiple-testing correction, ultimately generating a list of candidate risk genes. We defined genes with an FDR under 0.1 were considered “high-confidence” risk genes, a stringent threshold commonly used in similar rare variant studies to reduce false discoveries.^14,26–28^

### Differential expression analysis

Total RNA sequencing data of ADHD cases and controls were obtained from publicly available datasets on the Gene Expression Omnibus (GEO; http://www.ncbi.nlm.nih.gov/geo).^30^ We utilized ‘GSE159104’ study, which had whole-blood RNA-seq dataset composed of both monozygotic twin pairs (affected proband and unaffected sibling) and unrelated case-control samples, all characterized for ADHD status.^31^ Using the edgeR package,^85,86^ normalized gene expression matrix was analyzed to compare gene expression levels between 16 probands and their unaffected siblings, as well as between 23 ADHD cases and 21 controls. Gene-level statistical significance was determined using both unadjusted *P* values and multiple-testing corrected false discovery rates (FDR).

Additionally, to explore cell type-specific gene expression patterns in neural tissue, we leveraged single-cell RNA sequencing data accessed through the CELLxGENE platform (https://cellxgene.cziscience.com/gene-expression).^32^ All 15 ADHD risk genes were profiled across diverse neural cell populations, including neurons and glial cells, to provide insight into their potential roles in ADHD pathophysiology.

#### Pathway enrichment analysis

To identify biological pathways enriched in ADHD risk genes, we performed pathway enrichment analysis using FUMA GWAS.^33^ The input consisted of a list of risk genes identified from the rare variant study or GWAS. To annotate risk genes from the current ADHD GWAS,^11^ we used MAGMA (Multi-marker Analysis of Genomic Annotation), a tool used for annotating GWAS results to identify genes associated with traits or diseases.^34^ It links SNPs to genes by mapping variants within a defined window around each gene, typically 10 kb upstream and downstream. MAGMA then aggregates SNP-level association signals into gene-level scores using statistical models that account for linkage disequilibrium (LD). We utilized the gene location files and European LD reference panel from MAGMA website (https://ctg.cncr.nl/software/magma). We kept annotated genes with *P* value < 2.83E-06 (0.05/17,635 genes). The merged gene lists were mapped to predefined pathways from curated databases, including Reactome, KEGG, and Gene Ontology biological processes (MsigDB c5). A statistical test was employed to assess whether the genes were significantly overrepresented in any pathways compared to the background gene set. Pathways with adjusted *P* values below the significance threshold (FDR < 0.05, Benjamini–Hochberg correction) were considered enriched.

### Protein-protein interaction analysis

To examine the protein-protein interactions (PPI) among the identified ADHD risk genes, we utilized the Search Tool for the Retrieval of Interacting Genes/Proteins (STRING; http://string-db.org).^35^ STRING calculates an enrichment *P* value by comparing the observed number of edges (i.e., protein-protein interactions) to the number expected by chance, given the size and composition of the input gene set. Additionally, functional enrichment analyses of the resulting networks were subsequently performed within STRING, applying Benjamini-Hochberg corrections for multiple testing to identify significantly enriched biological processes, pathways, and protein classes. We first generated a PPI network using the subset of 15 ADHD risk genes identified from our rare variant analysis, and then performed a similar analysis on an expanded set of 57 ADHD-associated genes (including the 15 rare variant genes and an additional 42 genes identified through GWAS). Network construction and analysis were carried out using default STRING parameters, with confidence interaction scores ≥0.15.

### Ancestry Analysis

We analyzed the 15 genes associated with ADHD risk to evaluate whether the genetic architecture involving rare damaging variants differs across ancestries. This study utilized data from the gnomAD database (version 4.1.0) and the All of Us Research Program (version 7, individuals without ADHD) to examine ancestry-specific differences in the frequency and burden of rare genetic variants. We focused on three major ancestry groups: non-Finnish European (NFE; 622,057 individuals in gnomAD and 129,532 in All of Us), African (AFR; 37,545 individuals in gnomAD and 47,631 in All of Us), and admixed-American (AMR; 30,019 individuals in gnomAD and 44,329 in All of Us). For the ADHD cases, we included 3,756 NFE, 464 AFR, and 518 AMR samples from the All of Us Research Program.

Rare variants were defined as those with a minor allele frequency (MAF) < 0.01% in at least one population. Population-level frequencies of rare PTV, MisB, MisA and SYN variants were obtained from the merged gnomAD and All of Us datasets. For PTVs, we used LOFTEE, retaining variants labeled as “high-confidence” (HC) and excluding those flagged by LOFTEE, except for the “SINGLE_EXON” category.

Comparisons of variant burdens between populations were performed using Fisher’s exact test. Additionally, comparisons of ADHD cases and population-level data between ancestry groups were adjusted for baseline frequency differences using the Breslow–Day test for homogeneity of odds ratios. Statistical significance was defined as a P value below 0.001, applying Bonferroni correction for multiple testing (0.05/45).

### Relation between ADHD and ASD

To assess whether the burden of deleterious rare coding variants differed between ADHD and ASD, we utilized variant data from a large ASD whole-exome sequencing (WES) dataset comprising 15,036 ASD probands and 5,591 ASD cases.^25^ We first kept de novo and case-control variants in the 15 ADHD risk genes, identified in our rare variant analysis, from the ASD WES study. We then classified them into PTVs, MisB, and MisA and merged the counts of the deleterious variants. We also quantified the number of those deleterious variants within the 15 ADHD risk genes and compared these counts to those observed across all other genes in the exome (18,128 genes, excluding the 15 ADHD risk genes). To test for heterogeneity in variant enrichment between ADHD and ASD, we conducted Fisher’s exact tests, evaluating whether the observed frequencies of deleterious variants in ADHD-related genes differed significantly from those in ASD cohorts. Multiple testing corrections were performed using the Benjamini-Hochberg procedure.

## Acknowledgements

This study was supported by a grant from Beatrice and Samuel A. Seaver Foundation (BM); and the National Institutes of Health (NIH), under grant numbers R01MH129724 (BM), 5R01MH097849 (BM), and K08MH128665 (EO).

The All of Us Research Program is supported by the National Institutes of Health, Office of the Director: Regional Medical Centers: 1 OT2 OD026549; 1 OT2 OD026554; 1 OT2 OD026557; 1 OT2 OD026556; 1 OT2 OD026550; 1 OT2 OD 026552; 1 OT2 OD026553; 1 OT2 OD026548; 1 OT2 OD026551; 1 OT2 OD026555; IAA #: AOD 16037; Federally Qualified Health Centers: HHSN 263201600085U; Data and Research Center: 5 U2C OD023196; Biobank: 1 U24 OD023121; The Participant Center: U24 OD023176; Participant Technology Systems Center: 1 U24 OD023163; Communications and Engagement: 3 OT2 OD023205; 3 OT2 OD023206; and Community Partners: 1 OT2 OD025277; 3 OT2 OD025315; 1 OT2 OD025337; 1 OT2 OD025276. In addition, the All of Us Research Program would not be possible without the partnership of its participants.

## Author contributions

Study concept and design: MC, SJ, BM

Acquisition, analysis, or interpretation of data: MC, SJ, BM

Drafting of the manuscript: MC, TVF, SJ, BM, EO

Critical revision of the manuscript for important intellectual content: All authors

Statistical analysis: MC, SJ, BM

Obtained funding: BM

Study supervision: BM

## Competing interests

The authors declare no competing interests.

## Data availability

All of Us controlled tier data is available for registered institutions and cohort selection is detailed in the workbench titled “Detecting the prevalence of ultra-rare gene mutations.” Due to stringent privacy protections and data sharing restrictions enforced by the All of Us Research Program, individual-level variant data cannot be shared publicly. Researchers must adhere to specific guidelines to protect participant confidentiality, which includes not reporting or disseminating any data that could allow the re-identification of participants, particularly those involving participant counts between 1 and 20 without employing approved data obscuration methods.

Details of the Olfson *et al.* dataset are described in the following publication by Emily Olfson, Luis C. Farhat, Wenzhong Liu, Lawrence A. Vitulano, Gwyneth Zai, Monicke O. Lima, Justin Parent, Guilherme V. Polanczyk, Carolina Cappi, James L. Kennedy, and Thomas V. Fernandez: Rare de novo damaging DNA variants are enriched in attention-deficit/hyperactivity disorder and implicate risk genes. Sequence generation was supported by a Klingenstein Third Generation Foundation ADHD Fellowship grant (E.O.), a Yale Child Study Center Faculty Development Award (T.V.F.), and the Allison Family Foundation (T.V.F.).

## Code availability

Code and resources used in this study is available on GitHub at (https://github.com/MahjaniLab/ADHD_WES). For any further inquiries or requests for code not available in the repository, please contact the corresponding author.

## Notes

### Competing Interest Statement

The authors have declared no competing interest.

### Author Declarations

The use of de-identified data from the All of Us Research Program, accessed via the Researcher Workbench, does not constitute human subjects research and does not require IRB review according to the guidelines provided in the All of Us Responsible Conduct of Research training. Our project adheres to the ethical principles of research with human participants, as encouraged by the All of Us program, although it involves only the analysis of previously collected, de-identified data.

