## Extended data figures for "Rare Variant Analyses in Ancestrally Diverse Cohorts Reveal Novel ADHD Risk Genes"

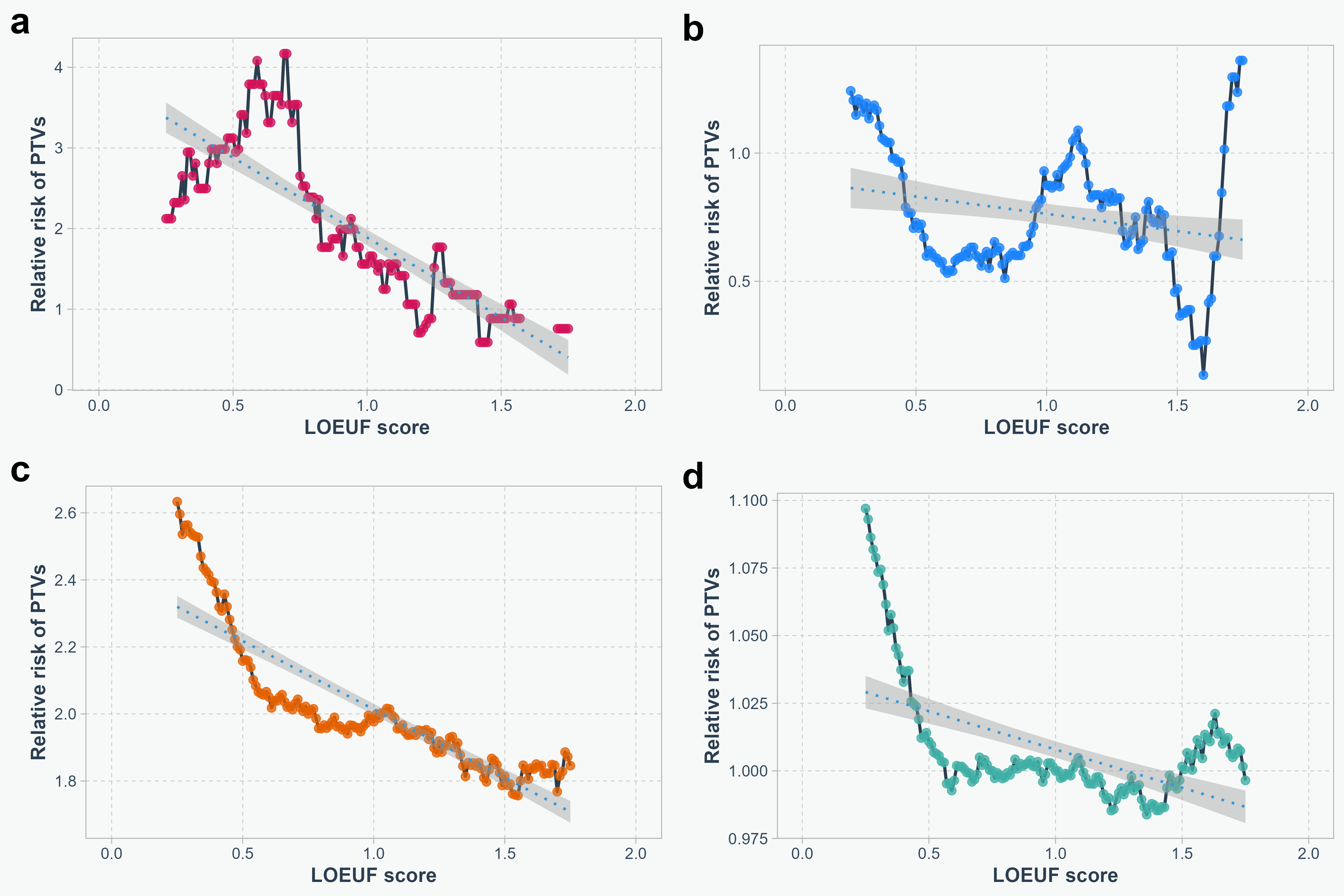

**Extended Data Fig. 1: Relative risk of protein-truncating variants across LOEUF scores in each study cohort.** Relative risk of protein-truncating variants (PTVs) as a function of the LOEUF (loss-of-function observed/expected upper bound fraction) score, calculated using genes within a sliding window of 25% of the LOEUF distribution. Each panel represents results for a different dataset or stratification: Relationship between relative risk of PTVs and LOEUF score in the (a) Olfson data, (b) SPARK data, (c) Satterstrom data, and (d) All of Us data. Dots represent the calculated relative risk in each window, solid lines denote smoothed trends, and dotted blue lines highlight overall linear trends across the data. Shaded regions indicate 95% confidence intervals for the linear regression model.

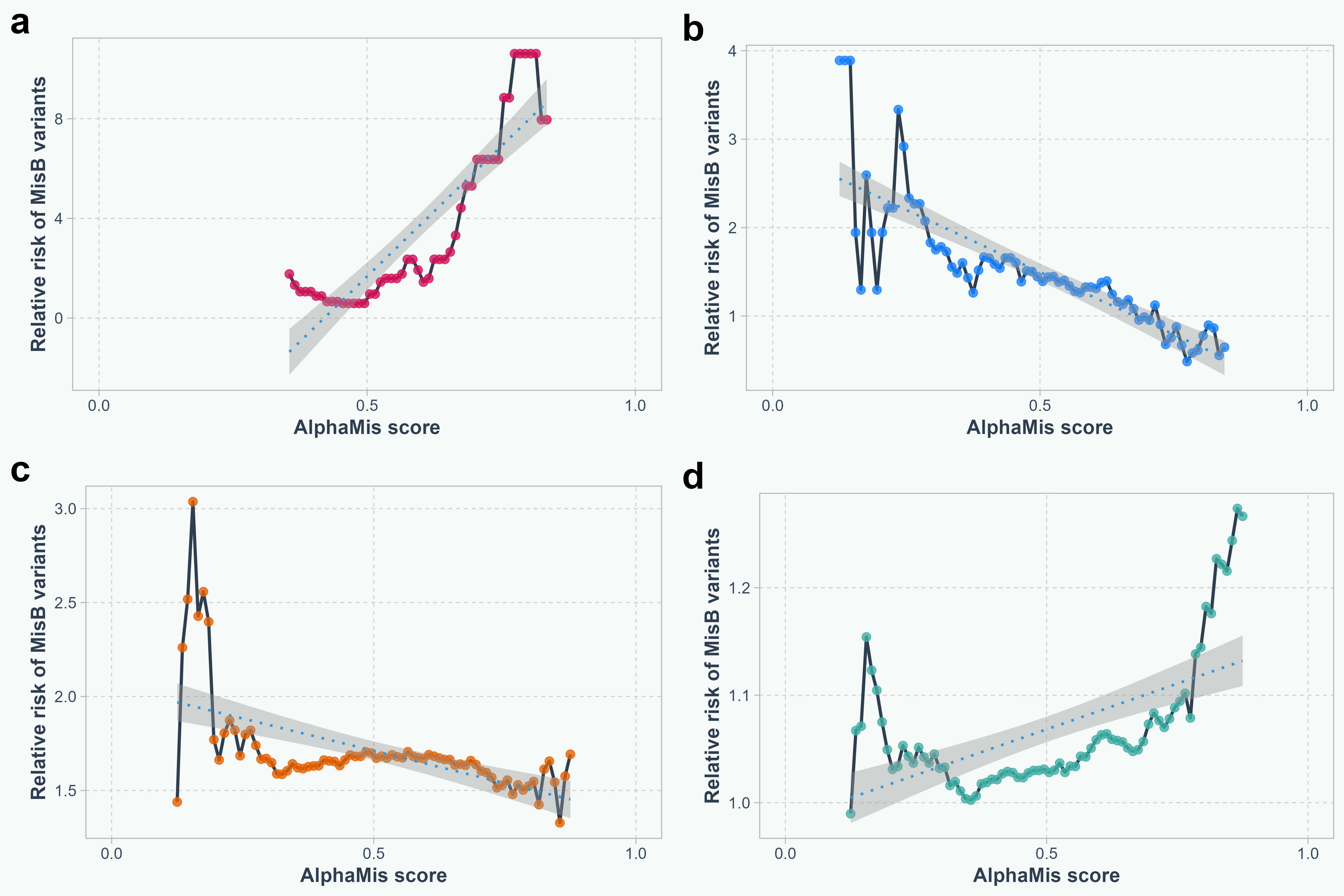

**Extended Data Fig. 2: Relative risk of MisB across AlphaMissense scores in each study cohort.** Relative risk of MisB, missense variants with MPC (Missense badness, PolyPhen-2, and Constraint) score ≥ 2, as a function of the AlphaMissense score, calculated using genes within a sliding window of 25% of the AlphaMissense distribution. Each panel represents results for a different dataset or stratification: Relationship between relative risk of MisB and AlphaMissense score in the (a) Olfson data, (b) SPARK data, (c) Satterstrom data, and (d) All of Us data. Dots represent the calculated relative risk in each window, solid lines denote smoothed trends, and dotted blue lines highlight overall linear trends across the data. Shaded regions indicate 95% confidence intervals for the linear regression model.

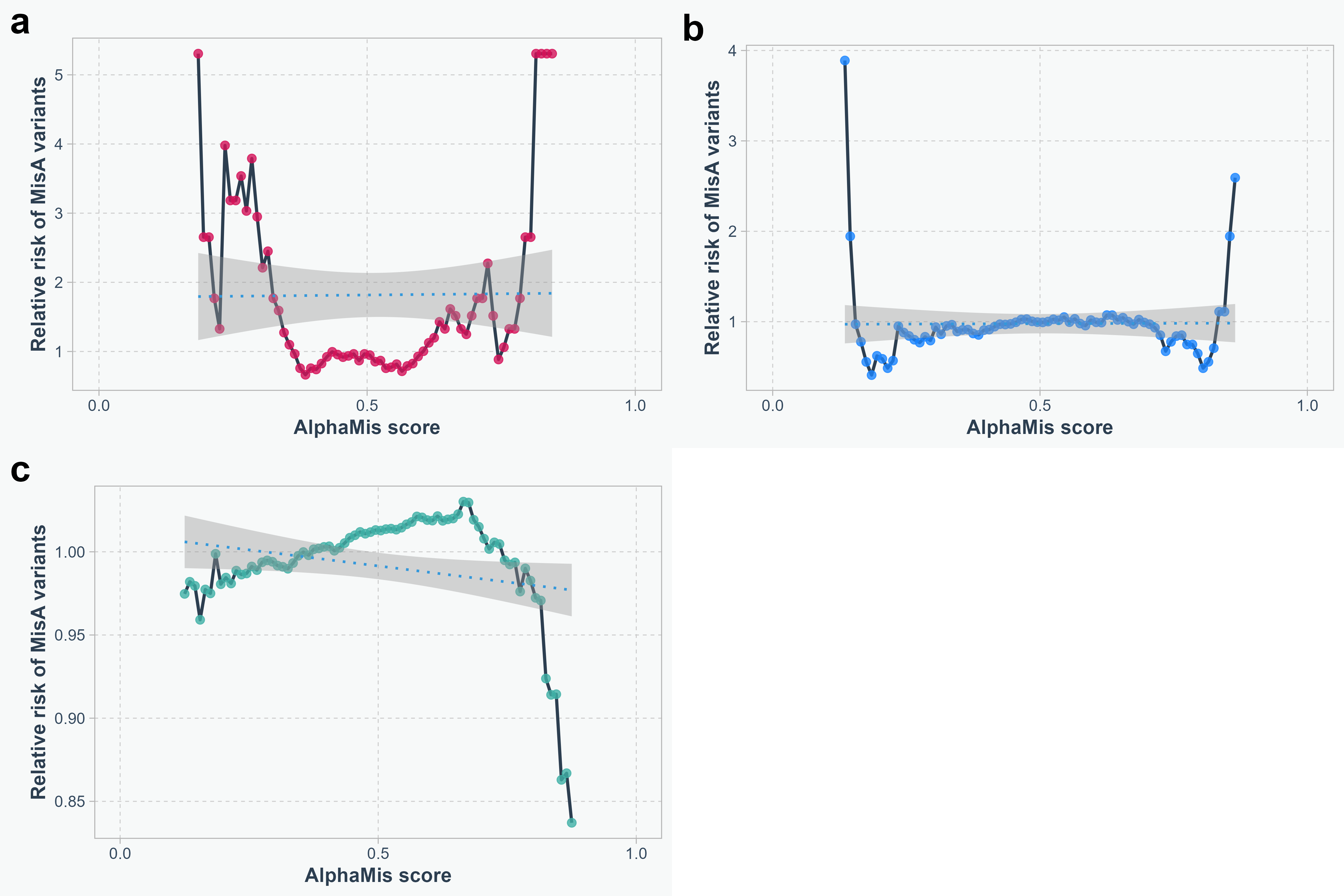

**Extended Data Fig. 3: Relative risk of MisA across AlphaMissense scores in each study cohort.** Relative risk of MisA, missense variants with 2 > MPC (Missense badness, PolyPhen-2, and Constraint) score ≥ 1, as a function of the AlphaMissense score, calculated using genes within a sliding window of 25% of the AlphaMissense distribution. Each panel represents results for a different dataset or stratification: Relationship between relative risk of MisA and AlphaMissense score in the (a) Olfson data, (b) SPARK data, and (c) All of Us data. MisA variants were not available in the Satterstrom dataset. Dots represent the calculated relative risk in each window, solid lines denote smoothed trends, and dotted blue lines highlight overall linear trends across the data. Shaded regions indicate 95% confidence intervals for the linear regression model.

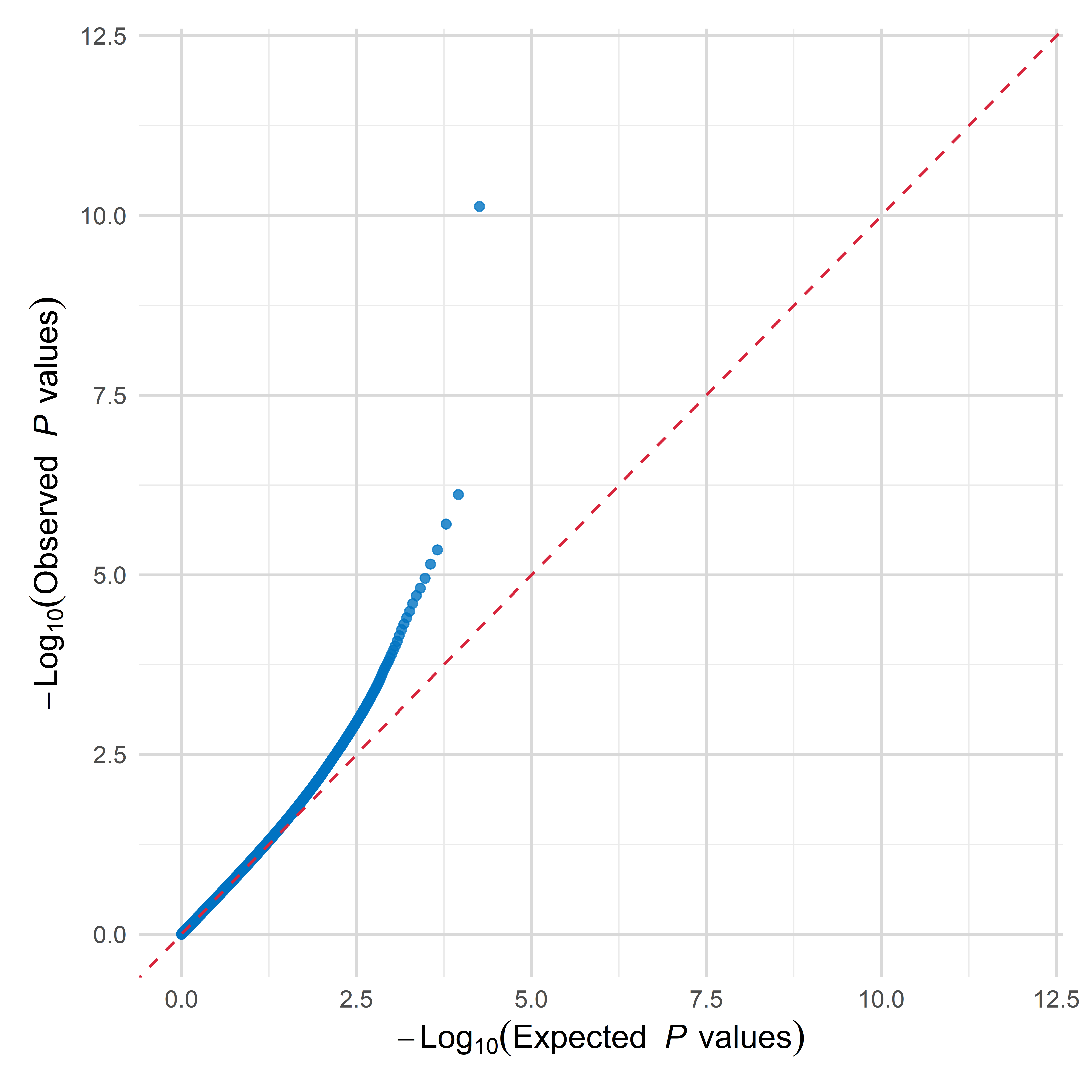

**Extended Data Fig. 4: Quantile-quantile plots of TADA (Transmitted and De Novo Association) analysis.** The plot compares the observed -log_10_(*P* values) against the expected -log_10_(*P* values) under the null hypothesis. The red diagonal line represents the expected distribution of *P* values if there is no association.

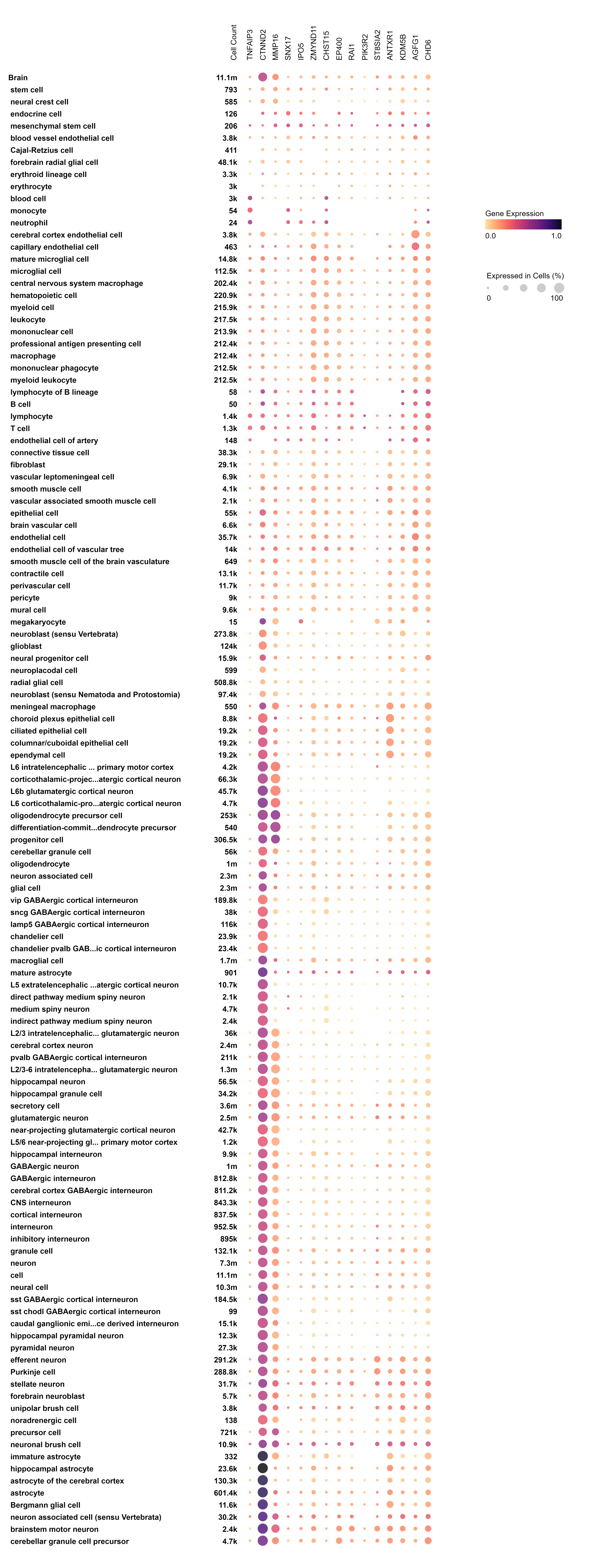

**Extended Data Fig. 5: Expression patterns of 15 ADHD risk genes across diverse cell types in the brain.** The x-axis represents the 15 risk genes for ADHD, annotated with their expression levels and distribution across distinct cell types. The y-axis lists cell types hierarchically, categorized by lineage, including cortical, stem, and immune cells. Dot size reflects the percentage of cells within each cell type expressing the gene, with larger dots indicating higher percentages. Dot color denotes the normalized average expression level, ranging from light (low expression) to dark purple (high expression). Cell counts for each cell type are displayed adjacent to the y-axis labels.

**a**

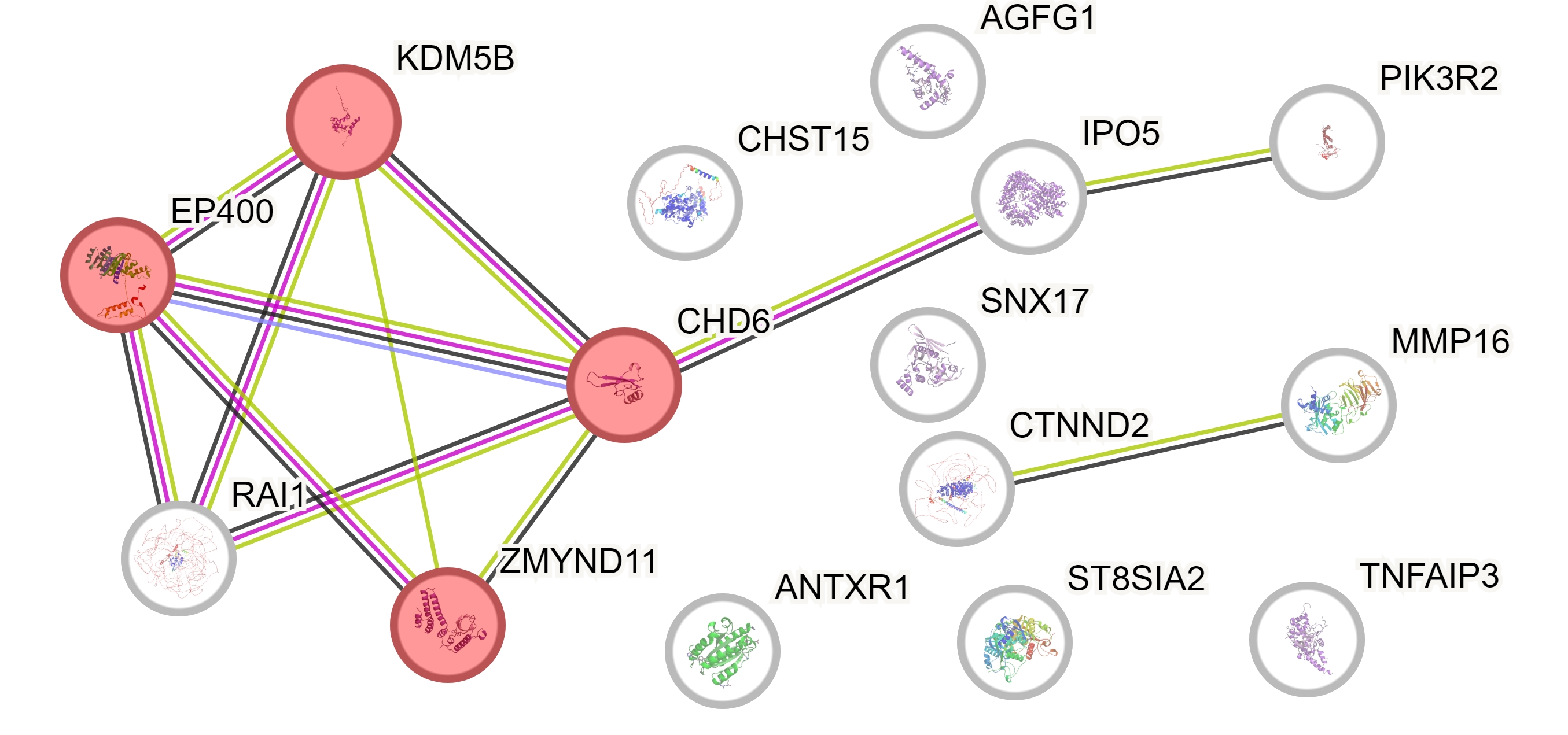

**b**

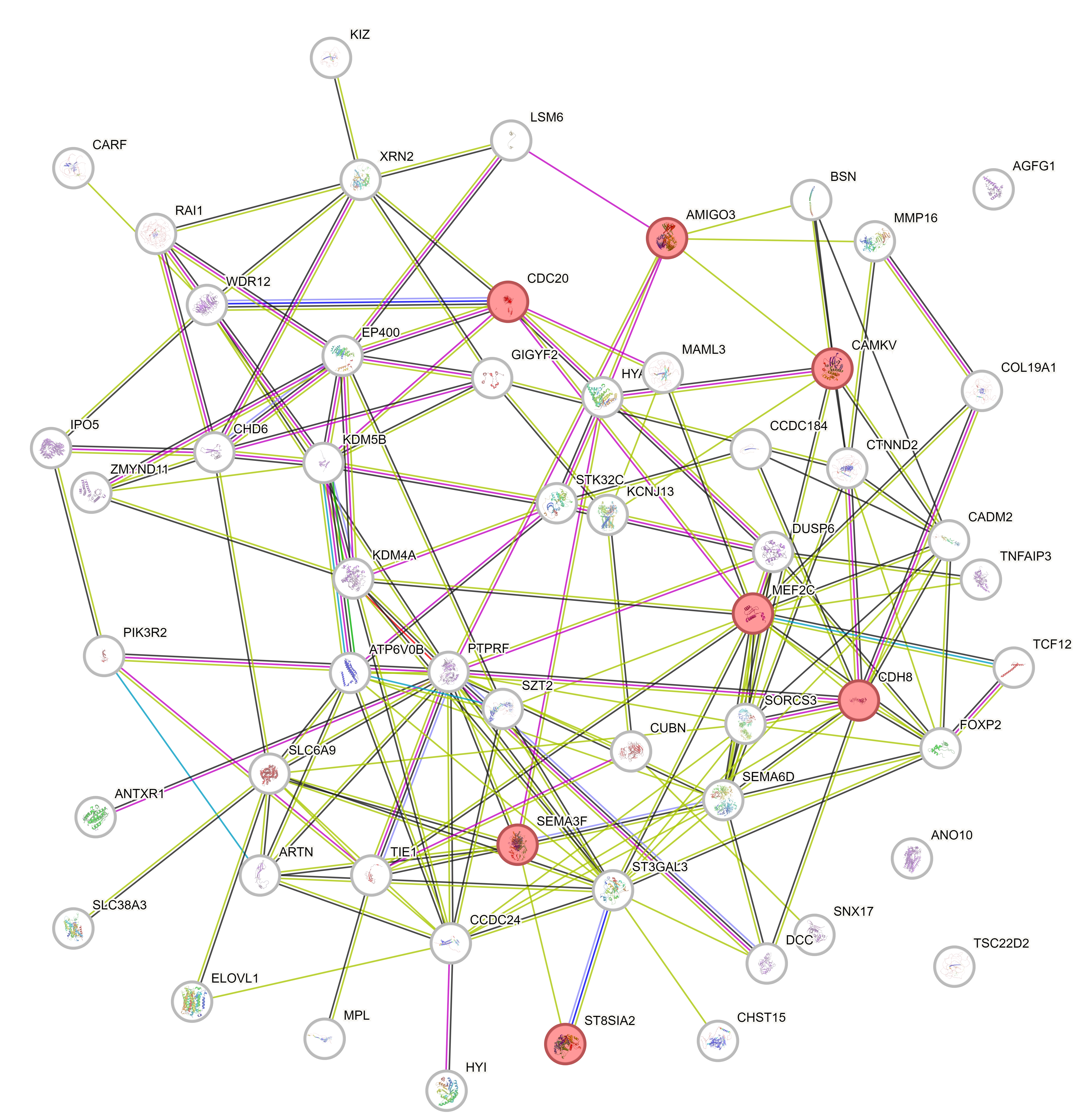

**Extended Data Fig. 6: Protein-protein interactions between ADHD risk genes.** Protein-protein interaction (PPI) networks of **a)** the 15 ADHD risk genes identified from the rare variant analysis and **b)** the combined set of 57 ADHD-associated genes (15 from the rare variant study and 42 from GWAS) were visualized, where each node represents a protein labeled with its corresponding gene name. The connections (edges) between nodes denote predicted or experimentally validated interactions. Red-colored nodes indicate proteins associated with ‘chromatin regulator’ in ‘**a**’, and ‘regulation of synapse organization’ in ‘**b**’. Edge colors signify different types of interaction evidence: light blue edges represent known interactions from curated databases, pink edges indicate experimentally determined interactions, blue edges show co-occurrence evidence, black edges denote co-expression-based evidence, green edges represent genome neighborhood-based interactions, light green edges signify evidence from text mining, and light purple edges indicate protein homology-based associations. Each node includes a visual depiction of the protein structure, highlighting its known or predicted conformation.

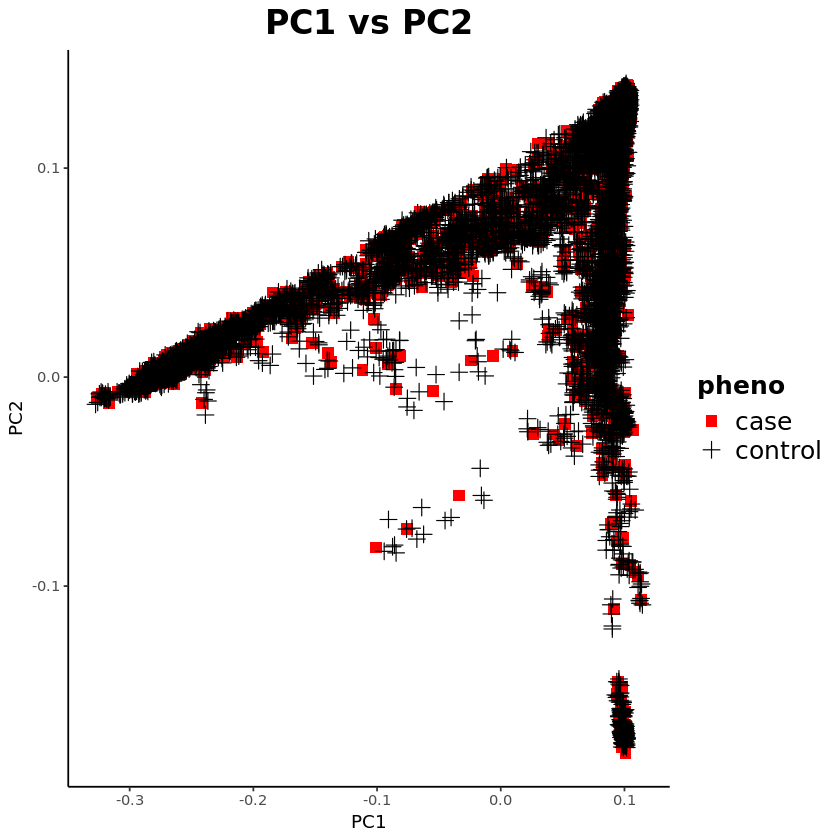

**Extended Data Fig. 7: The principal component analysis (PCA) plot of 5,063 ADHD cases and 25,315 controls in the All of Us Research Program.** The first principal component (PC1) is on the x-axis and the second principal component (PC2) is on the y-axis. Each point represents an individual, color-coded by predicted population and shaped by phenotype status (case or control).
